# Reopening universities during the COVID-19 pandemic: A testing strategy to minimize active cases and delay outbreaks

**DOI:** 10.1101/2020.07.06.20147272

**Authors:** Lior Rennert, Corey A. Kalbaugh, Lu Shi, Christopher McMahan

## Abstract

**Background:** University campuses present an ideal environment for viral spread and are therefore at extreme risk of serving as a hotbed for a COVID-19 outbreak. While active surveillance throughout the semester such as widespread testing, contact tracing, and case isolation, may assist in detecting and preventing early outbreaks, these strategies will not be sufficient should a larger outbreak occur. It is therefore necessary to limit the initial number of active cases at the start of the semester. We examine the impact of pre-semester NAT testing on disease spread in a university setting.

**Methods:** We implement simple dynamic transmission models of SARS-CoV-2 infection to explore the effects of pre-semester testing strategies on the number of active infections and occupied isolation beds throughout the semester. We assume an infectious period of 3 days and vary *R*_*0*_ to represent the effectiveness of disease mitigation strategies throughout the semester. We assume the prevalence of active cases at the beginning of the semester is 5%. The sensitivity of the NAT test is set at 90%.

**Results:** If no pre-semester screening is mandated, the peak number of active infections occurs in under 10 days and the size of the peak is substantial, ranging from 5,000 active infections when effective mitigation strategies (*R*_*0*_ = 1.25) are implemented to over 15,000 active infections for less effective strategies (*R*_*0*_ = 3). When one NAT test is mandated within one week of campus arrival, effective (*R*_*0*_ = 1.25) and less effective (*R*_*0*_ = 3) mitigation strategies delay the onset of the peak to 40 days and 17 days, respectively, and result in peak size ranging from 1,000 to over 15,000 active infections. When two NAT tests are mandated, effective (*R*_*0*_ = 1.25) and less effective (*R*_*0*_ = 3) mitigation strategies delay the onset of the peak through the end of fall semester and 20 days, respectively, and result in peak size ranging from less than 1,000 to over 15,000 active infections. If maximum occupancy of isolation beds is set to 2% of the student population, then isolation beds would only be available for a range of 1 in 2 confirmed cases (*R*_*0*_ = 1.25) to 1 in 40 confirmed cases (*R*_*0*_ = 3) before maximum occupancy is reached.

**Conclusion:** Even with highly effective mitigation strategies throughout the semester, inadequate pre-semester testing will lead to early and large surges of the disease and result in universities quickly reaching their isolation bed capacity. We therefore recommend NAT testing within one week of campus return. While this strategy is sufficient for delaying the timing of the outbreak, pre-semester testing would need to be implemented in conjunction with effective mitigation strategies to reduce the outbreak size.

## Introduction

Universities in the United States are currently exploring strategies to mitigate the spread of COVID-19 prior to their planned reopening in fall 2020. Between dormitories, classrooms, and nightlife, university campuses present an ideal environment for viral spread and are therefore at extreme risk of serving as a hotbed for a COVID-19 outbreak. Many universities have widespread testing capabilities, contact tracing, and reserved spaces to isolate detected cases, which are well-known strategies for controlling COVID-19 in the absence of a vaccine.^1–4^ Recent modeling studies found that widespread testing of the entire student population once per month was sufficient for detecting an outbreak of fewer than 9 individuals,^3^ and small outbreaks could be contained with highly effective contact tracing and case isolation.^4^

However, these strategies may not be sufficient for curtailing a larger outbreak should one occur. This is primarily due to the number of daily high-density social events, especially during the first few weeks of the semester, that occur on and off campus. These events lead to a high number of close-encounter contacts per student and thus provides a perfect path for viral transmission. Because cases are not detected immediately, it will be virtually impossible to trace all close-encounter contacts of infected individuals. A recent study concluded that contact tracing and isolation are likely ineffective in controlling outbreaks if the number of initial cases is 40 or greater.^4^ Given the difficulties of timely contact tracing in the university setting, it is of utmost importance to limit the initial number of active cases at the start of the semester. One method of doing so is through student testing prior to their campus return.

There are differing views as to whether universities should test all students for COVID-19 prior to campus arrival. While some public officials stress the need to test in order to contain the spread of the virus, others cite the costs and false negatives of tests as reasons to limit testing and isolation to symptomatic students upon their return to campus.^5^ Currently, the CDC does not overtly endorse, nor recommend against, testing the entire student population prior to campus arrival.^6^ However, the impact of pre-semester testing strategies have not been previously studied.^6^

Our team was tasked with recommending strategies for baseline testing and isolation bed capacity to be used in moving a large university in the Southeastern United States toward re-opening amidst the ongoing COVID-19 pandemic. We considered several testing strategies: a) No screening of students prior to the fall semester, b) requiring all students to present a negative test (via a nucleic acid amplification test: NAT) within one week of arrival, and c) requiring all students to present two negative NAT tests within one week of arrival on campus.

## Methods

To guide and inform our recommendations, we implemented a simple dynamic transmission model^7^ of SARS-CoV-2 infection under each testing strategy to determine the number of active infections, occupied isolation beds, and days until the maximum isolation bed occupancy is reached. The latter could be viewed as a potential “trigger” that would initiate the closure of the university. We set N = S + I + R, where S is the number of susceptible individuals, I is the number of infected individuals, and R is the number of recovered individuals.^7^ In all scenarios, we set N = 25,000 and assume that 10% of students^3^ will have already had the disease and recovered by the start of the semester. We set the sensitivity of each NAT test at 90%.^8^ Table 1 presents the percentage of active cases at the beginning of the semester for each testing strategy, assuming different levels of active infection rates in the student population.

**Table 1:** Percentage (and expected number) of active cases under different initial active infection rates and testing strategies. The (expected) number of active cases are based on a university of size *N* = 25,000 and assumes that 10% of the population is immune (e.g., through previous disease exposure). NAT test sensitivity is set at 90%.

| % infectious at beginning of semester | % of student population infectious on campus for each testing strategy (expected number of active cases on campus for $N=25,000$) | | |
| --- | --- | --- | --- |
|  | No testing | One NAT test | Two NAT tests |
| 1% | 1% (225 cases) | 0.1% (23 cases) | 0.01% (2 cases) |
| 5% | 5% (1125 cases) | 0.5% (113 cases) | 0.05% (11 cases) |
| 10% | 10% (2250 cases) | 1% (225 cases) | 0.1% (23 cases) |

We assume an initial active infection rate of 5% for the transmission model. This number is based on the current number of active infections in South Carolina^9^, multiplied by a factor of ten to capture underreporting.^10^ For each testing strategy, the number of infected individuals on campus at the start of the semester are provided in the second row of Table 1. In the models below, we specify the maximum occupancy of isolation beds to be 2% of the student population. For a university with a student population of 25,000, this is equivalent to 500 isolation beds. We assume that students remain in isolation for 11 days, which represents the median time until a negative NAT test.^11^

The average infectious period, 1/γ, is set to 3 days. We vary the level of *R*_*0*_ between 1.25, 1.5, 2, and 3 during the infectious period. The Center for Disease Control and Prevention (CDC) estimates *R*_*0*_ in the range of 2 to 3.^12^ The lower values of *R*_*0*_ imply a reduced contact rate between students and reflect effective implementation of mitigation strategies, such as successful contact tracing and isolation of suspected and confirmed cases, enforcement of social distancing, mask mandates, etc. For a fixed *R*_*0*_, the contact rate multiplied by the probability of transmission given contact between a susceptible and infectious individual is given by β = *R*_*0*_ · γ.^7^ These models assume a closed epidemic in which no new entries or deaths are allowed.

## Results

If no pre-semester screening is mandated, the peak number of active infections occurs in 10 days (Figure 1). Furthermore, the size of the peak is substantial, ranging from 5,000 active infections when effective mitigation strategies are implemented (*R*_*0*_ = 1.25) to over 15,000 active infections for less effective strategies (*R*_*0*_ = 3). Requiring at least one NAT test within one week of campus arrival delays the peak number of active infections and the corresponding time until isolation bed occupancy is reached. The advantages of pre-semester screening are especially noteworthy when implemented in conjunction with effective mitigation strategies (as indicated by the lower values of *R*_*0*_).

**Figure 1.**
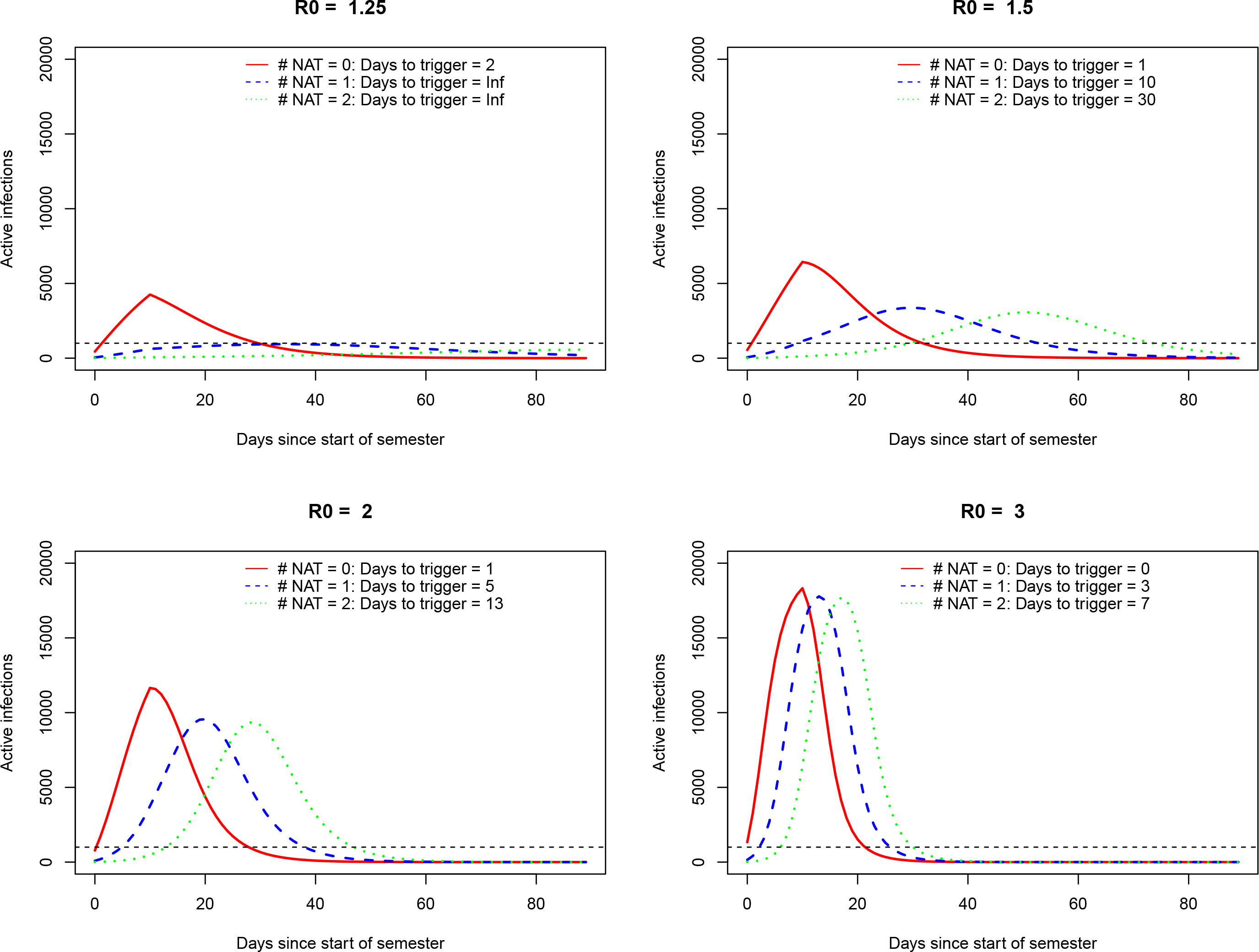
Expected number of active infections over time under three pre-semester testing strategies: No NAT tests (solid red line), 1 NAT test (dashed blue line), 2 NAT tests (dotted green line). Assuming 50% of infected students are given an isolation bed and a total of 500 isolation beds are available, the trigger will be reached when there are 1000 active infections (on average). This threshold is represented by the horizontal dashed line.

When one NAT test is mandated within one week of campus arrival, effective (*R*_*0*_ = 1.25) and less effective (*R*_*0*_ = 3) mitigation strategies delay the onset of the peak to 40 days and 17 days, respectively, and result in peak size ranging from 1,000 to over 15,000 active infections. When two NAT tests are mandated, effective (*R*_*0*_ = 1.25) and less effective (*R*_*0*_ = 3) mitigation strategies delay the onset of the peak through the end of fall semester and 20 days, respectively, and result in peak size ranging from less than 1,000 to over 15,000 active infections.

However, unless university mitigation strategies are indeed effective, maximum isolation bed occupancy is reached quickly. If isolation beds are occupied by 50% of confirmed cases, then pre-semester screening will not maintain isolation bed occupancy below the maximum threshold for values of *R*_*0*_ greater than 1.25. If values of *R*_*0*_ are in the 2 to 3 range, then isolation beds would only be available for a range of 1 in 20 to 1 in 40 confirmed cases before maximum occupancy is reached.

## Discussion

The results of our modeling study highlight the importance of detecting active cases prior to campus arrival. This is essential to ensuring that a rapid spike of cases does not occur in the beginning of the semester; a failure to do so can result in bringing anywhere from hundreds to thousands of infectious cases on campus. Our team has therefore recommended that each student be tested (via NAT) at least once within one week of returning to campus. Ideally, given the biology of the virus and the operating characteristics of the various NAT tests available for COVID-19, we believe that each student should be tested twice within a 3-day interval during this period to minimize the risk of false negatives.^13^ However, even with pre-semester screening, highly effective mitigation strategies are needed to avoid a large surge in cases. This will result in universities quickly reaching their isolation bed capacity and may force healthy students to live with infectious roommates. Universities must therefore give careful consideration to isolation strategies for suspected or confirmed cases.

## Limitations

There are several limitations of the proposed strategies. It remains unclear as to whether insurance providers will cover the costs of precautionary tests.^14^ Therefore the financial burden may fall on the students or the university. In addition, single administration of a test could miss cases in the early stages of infection, as well as cases that occur in the days between adminstration of the test and campus arrival.^6^ For these reasons, a second NAT test has been recommended. However, this strategy amplifies the financial concerns and introduces additional logistical challenges. Universities may need to implement the second test upon arrival on campus and thus need to ensure that proper resources are in place.^6^

There is also uncertainty in the model parameters informing our study. However, we have performed sensitivity analyses by examining varying levels of reproductive rates. More robust data on test sensitivity, disease prevalence at the onset of the fall semester, and impact of mitigation strategies on disease spread would vastly improve our estimates by minimizing uncertainty.

## Conclusion

Detection of SARS-COV-2 prior to campus arrival is necessary to avoid a large outbreak of hundreds to thousands of active infections at the onset of the fall semester. This is achievable through pre-semester screening via NAT testing of the entire student population prior to campus arrival. While intensive pre-semester testing will delay the outbreak time, the size of the outbreak will only be reduced if highly effective mitigation strategies are implemented throughout the semester.

## Data Availability

Data sources guiding choice of all model parameters provided in manuscript. Code for transmission models will be made available on github.

